# Impact of BNT162b2 vaccination and isolation on SARS-CoV-2 transmission in Israeli households: an observational study

**DOI:** 10.1101/2021.07.12.21260377

**Authors:** Maylis Layan, Mayan Gilboa, Tal Gonen, Miki Goldenfeld, Lilac Meltzer, Alessio Andronico, Nathanaël Hozé, Simon Cauchemez, Gili Regev-Yochay

**Affiliations:** Mathematical Modelling of Infectious Diseases Unit, Institut Pasteur, UMR2000, CNRS, Paris, France; Sorbonne Université, Paris, France; Infectious Disease Unit, Sheba Medical Centre, Ramat-Gan, Israel; Sackler School of Medicine, Tel Aviv University, Tel Aviv, Israel; Infection Prevention & Control Unit, Sheba Medical Center, Ramat-Gan, Israel

## Abstract

**Background:** Massive vaccination rollouts against SARS-CoV-2 infections have facilitated the easing of control measures in countries like Israel. While several studies have characterized the effectiveness of vaccines against severe forms of COVID-19 or SARS-CoV-2 infection, estimates of their impact on transmissibility remain limited. Here, we evaluated the role of vaccination and isolation on SARS-CoV-2 transmission within Israeli households.

**Methods:** From December 2020 to April 2021, confirmed cases were identified among healthcare workers of the Sheba Medical Centre and their family members. Households were recruited and followed up with repeated PCR for a minimum of ten days after case confirmation. Symptoms and vaccination information were collected at the end of follow-up. We developed a data augmentation Bayesian framework to ascertain how age, isolation and BNT162b2 vaccination with more than 7 days after the 2^nd^ dose impacted household transmission of SARS-CoV-2.

**Findings:** 210 households with 215 index cases were enrolled. 269 out of 687 (39%) household contacts developed a SARS-CoV-2 infection. Of those, 170 (63%) developed symptoms. Children below 12 years old were less susceptible than adults/teenagers (Relative Risk RR=0·50, 95% Credible Interval CI 0·32-0·79). Vaccination reduced the risk of infection among adults/teenagers (RR=0·19, 95% CI 0·07-0·40). Isolation reduced the risk of infection of unvaccinated adult/teenager (RR=0·11, 95% CI 0·05-0·19) and child contacts (RR=0·16, 95% CI 0·07-0·31) compared to unvaccinated adults/teenagers that did not isolate. Infectivity was significantly reduced in vaccinated cases (RR=0·22, 95% CI 0·06-0·70).

**Interpretation:** Within households, vaccination reduces both the risk of infection and of transmission if infected. When contacts were not vaccinated, isolation also led to important reductions in the risk of transmission. Vaccinated contacts might reduce their risk of infection if they isolate, although this requires confirmation with additional data.

**Funding:** Sheba Medical Center.

**Research in context:** *Evidence before this study:* The efficacy of vaccination against severe acute respiratory syndrome coronavirus 2 (SARS-CoV-2) transmissions in households remains understudied. On June 28, 2021, we searched PubMed and medRxiv for articles published between December 1, 2020, and June 28, 2021, using the following combination of search terms: (“COVID-19” OR “SARS-CoV-2”) AND (“household*” OR “famil*”) AND “transmission” AND “vaccination”. Our search yielded two articles that investigated the effect of vaccination on SARS-CoV-2 transmission in households. They showed a lower risk of infection in households with vaccinees. Vaccine efficacy on the risk of infection was estimated to 80% after the 2^nd^ dose, and vaccine efficacy on the risk of transmission if infected was estimated to 49% 21 days after the 1^st^ dose. However, these estimates are derived from surveillance data with no active follow-up of the households. In addition, the impact of isolation precautions has not been assessed.

*Added value of this study:* Based on the active follow-up of households of health care workers from the Sheba Medical Center in Israel, we estimated the effect of vaccination on household transmission. To our knowledge, our study is the first to conjointly investigate the effect of vaccination, age, and isolation precautions on the risk of infection and the risk of transmission in households while accounting for tertiary infections in the household, infections within the community, the reduced infectivity of asymptomatic cases, misidentification of index cases, and household size. Our study confirmed the high efficacy of BNT-162b2 vaccination to reduce infection risk and transmission risk. It also suggests that isolation might remain beneficial to vaccinated contacts.

*Implications of all the available evidence:* Vaccination reduces susceptibility to infection and case infectivity in households. Isolation precautions also mitigate the risk of infection and should be implemented whenever a household member is infected. They might remain beneficial to vaccinated contacts.

## Introduction

Severe acute respiratory syndrome coronavirus 2 (SARS-CoV-2) is a highly transmissible virus that was first detected in Wuhan China in December 2019.^1,2^ It is the cause of coronavirus disease 2019 (COVID-19), that has spread through the world, leading to a pandemic that has infected at least 180M people and caused more than 3·9M deaths worldwide by July 1, 2021.^3^ The advent of novel COVID-19 vaccines has been an important breakthrough in the management of the pandemic. To determine how vaccination may modify epidemic dynamics, it is essential to estimate its effectiveness with respect to infection, transmission, and disease severity. Multiple studies have shown that SARS-CoV-2 vaccines are very effective at reducing both the risk of infection,^4–7^ and the risk of developing severe symptoms.^4,8,9^

Data documenting vaccine impact on transmission are more limited.^10–12^ This stems from the difficulty to thoroughly document chains of transmission and account for how different types of contacts may lead to different risks of transmission.^13^ Households represent the perfect environment to evaluate factors impacting transmission such as vaccination because the probability of SARS-CoV-2 transmission among household members is high, ranging between 14 and 32%.^14–16^ Beyond the evaluation of vaccine efficacy, understanding how vaccines affect household transmission is also important to determine how recommendations should evolve with vaccines. For example, should isolation precautions be maintained in partially vaccinated households? ^17^ Harris et al^11^ estimated vaccine impact on SARS-CoV-2 household transmission from large UK administrative datasets. However, they did not evaluate how isolation impacted the outcome and the passive nature of surveillance may have led to underestimating household transmission rates.

During the first months of 2021, Israel underwent its third wave due to the rise of the alpha variant representing approximatively 90% of the transmission. Concomittantly, vaccination was extended to all adults above 16 years old making Israel one of the first countries to reach high vaccination coverage of their population, with 60% of the total population being vaccinated by March 22, 2021.^3^ During this period, we followed up SARS-CoV-2 transmission in the households of 12,518 healthcare workers (HCW) of the Sheba Medical Center, the largest medical center in Israel. Here, we describe dynamics of transmission in these households and evaluate the impact vaccination and isolation measures had on these dynamics.

## Methods

### Study Design and Study Population

All HCW, regardless of vaccination status, were required to report daily by an electronic questionnaire on any COVID-19 related symptom they, or a member of their household, suffered. SARS-CoV-2 RT-qPCR was readily available and HCW were encouraged to be tested for any mild symptom or suspected exposures. All HCW were instructed to notify the infection prevention and control unit if one of their household members was SARS-CoV-2 positive. All SARS-CoV-2 detected HCW as well as those with a positive SARS-CoV-2 household member were immediately contacted as part of the epidemiologic investigation for contact tracing and were provided with instructions regarding isolation precautions. All unvaccinated household members were required to perform a PCR test on day one and ten after diagnosis of the positive COVID-19 patient. Vaccinated HCW were instructed to perform a PCR test every day they reported to the hospital for work and at least twice within the ten days after detection. Vaccinated household members were encouraged to perform two PCR tests during the ten days after detection.

Between December 31, 2020, and April 26, 2021, the HCW who were SARS-CoV-2 positive or reported a positive household member were contacted at least ten days after detection and were offered to join the study. Those who agreed and gave their consent, answered a telephone interview.

### Data and Sample Collection

Data collected during the phone interview included: the age and gender of their household members, their vaccination status, prior COVID-19 infections, their COVID-19 PCR test dates and results, their symptoms (i.e., fever, cough, myalgia, headache, congestion, diarrhea, vomiting, anosmia, or ageusia), the number of rooms and bathrooms in the household and the degree to which isolation precautions were adhered to. At the time of the study, only individuals 16 years old or older were eligible for vaccination.

The household member who had the first positive PCR test was defined as the index case. When multiple household members had a positive PCR test on the same day, they were defined as co-primary cases. We defined complete isolation as complete separation in sleeping and eating between household contacts and index case(s), and if a separate bathroom was provided for the index case(s). Partial isolation was defined if one of the above was violated but masks were continuously used, and eating was consistently separate.

For HCW nasopharyngeal swabs were collected by trained personnel and RT-qPCR was performed using the Allplex™ 2019-nCov RT-qPCR assay (Seegene Inc., S. Korea) and expressed by cycle threshold (Ct). Other household members reported the results of their COVID-19 test performed by their health care providers. Among the 693 household contacts, 547 had at least two RT-qPCR results, 71 had a single negative test at inclusion, and 75 did not perform any test during the follow-up period.

### Clinical outcome

Confirmed SARS-CoV-2 infections were defined by a positive RT-qPCR test, i.e., with a Ct value lower than 40. Symptomatic cases were defined as confirmed cases with the presence of at least one symptom among the following ones: fever, cough, myalgia, headache, congestion, diarrhea, vomiting, anosmia or ageusia. Contacts who reported at least one of the above-mentioned symptoms but were not confirmed because they performed no PCR test (n=6) or a single one at inclusion (n=2) were also considered as symptomatic cases. Asymptomatic cases were defined as confirmed cases who did not report any symptom over the follow-up period of the household.

### Statistical analysis

Baseline characteristics of the index cases and household contacts were described according to their vaccination status. All individuals above years 12 old are considered as adults/teenagers. The overall SAR was defined as the percentage of secondary cases among household contacts. We explored the effect of the type of contact on the SAR by calculating the SAR for different categories of household contacts: not isolated nor vaccinated adults/teenagers, vaccinated but not isolated adults/teenagers, isolated but not vaccinated adults/teenagers, vaccinated and isolated adults/teenagers, not isolated children, and isolated children. Here, isolation corresponds to complete or partial isolation. For each category, we defined the SAR as the proportion of secondary cases among household contacts belonging to the category. We also defined the SAR of vaccinated and unvaccinated index cases as the proportion of secondary cases in households with vaccinated or unvaccinated index cases, respectively. In a sensitivity analysis, the SAR calculation was restricted to households in which a single index case was identified.

We developed a statistical model to evaluate the effect of age, isolation precautions, BNT162b2 vaccination and household size on SARS-CoV-2 transmission dynamics in households. The model uses the sequence of symptom onset dates and positive molecular test dates to estimate the person-to-person risk of transmission within the household while accounting for the community hazard of infection (i.e., household contacts infected outside the household) and the possibility of tertiary transmissions (i.e., household contacts infected by a member of the household that is not the index case).^18^ The person-to-person risk of transmission is decomposed into the baseline person-to-person risk of infection that depends on household size, the relative infectivity of the infector that depends on their vaccination status (reference group: unvaccinated cases), and the relative susceptibility of the infectee that depends on their age, isolation behavior, and vaccination status. The relative susceptibility is estimated separately for not isolated children, isolated children, isolated and unvaccinated adults/teenagers, vaccinated and not isolated adults/teenagers, and adults/ teenagers that are both isolated and vaccinated, considering the group of adults/teenagers that are not vaccinated and do not isolate as the reference group. None of the children were vaccinated at the time of the study. This formulation accommodates for the potential confounding effects between the three variables characterizing household contacts. We assumed that individuals whose isolation behavior was missing (n=6) did not isolate from the index case.

Model parameters were estimated using a Bayesian Markov Chain Monte Carlo sampling with data augmentation.^18^ Data were augmented with the probable date of infection of confirmed cases. For symptomatic cases, the date of infection was reconstructed from the date of symptom onset, using the probabilistic distribution of the incubation period.^19^ For asymptomatic cases, we assumed that the date of infection could occur up to ten days prior to their molecular detection based on a meta-analysis.^20^

Since the study was conducted during the vaccine rollout, participants were enrolled at varying stages of their vaccination process. We assumed that vaccines reach their full effect 7 days after receiving their 2^nd^ dose^4,8,9^. Cases were therefore considered vaccinated if their infection occurred >7 days after the 2^nd^ dose. Similarly, household contacts were considered vaccinated if their exposure to the index case occurred >7 days after the 2^nd^ dose. In a sensitivity analysis, we investigated how parameter estimates changed under the assumption that vaccination is effective >15 days after the 1^st^ dose. We also assessed how estimates changed when the analysis was restricted to households in which all negative contacts had performed a PCR test in the ten days following the detection of the index case. In the baseline scenario, we assumed that asymptomatic cases are 40% less infectious than symptomatic cases based on a meta-analysis,^21^ and investigated whether assuming the same level of infectivity in asymptomatic and symptomatic cases modified our estimates. Finally, in our baseline analysis, we chose a log-normal with log-mean=0 and log-sd=1 prior distribution for the relative infectivity and relative susceptibility parameters and modified it (log-sd=0·7, 2) in a sensitivity analysis.

We compared the observed and expected distributions of the number of cases per household size to validate the goodness-of-fit of the model (see the Supplementary Materials). We report the posterior median and the 95% Credible Interval (CI) of estimated parameters. We also report the posterior probability that isolated and vaccinated adult/teenager contacts are less susceptible than vaccinated adult/teenager contacts that do not isolate. To measure the strength of evidence of a reduced susceptibility in isolated contacts among vaccinated ones, we report the associated Bayes Factor (BF). Here, it directly corresponds to the posterior odds of a reduced susceptibility in isolated contacts among vaccinated ones. Additional details are available in Supplementary Materials.

### Ethics

The study was approved by the Sheba Medical Center IRB committee (approval #8130-21).

## Results

All 12,518 HCWs employed by Sheba Medical Center were eligible to join the study. Between December 19 and April 28, 2021, 91% of Sheba Medical Center personnel received both doses of the BNT162b2 vaccine, and a rapid and significant decrease in newly detected cases within HCW was observed.

From December 31, 2020, to April 26, 2021, 276 SARS-CoV-2 cases were identified among HCWs of the Sheba Medical Center and their household members (Figure 1). Of these, 212 agreed to participate, gave their consent, and were enrolled in the study with their household members. Two households were excluded due to missing vaccination status, dates of PCR test and/or symptom onset. In total, we analyzed data from 210 households with 215 index cases, including 4 co-primary cases, and their 687 household contacts. The median household size was 4 (IQR 3-5). Mean age was 32 years in index cases and 27 years among household contacts (Table 1). Age was missing for 5 adult/teenager contacts and isolation behavior was missing for 6 contacts. There was a slight over-representation of females among index cases (58%). 191 index cases (89%) were adults/teenagers, of whom 15 (8%) were vaccinated. None of the 24 child index cases were vaccinated. Among the 494 adult/teenager household contacts, 124 (25%) were vaccinated. Of these, 83 (17%) also isolated. Among the 370 unvaccinated adult/teenager contacts, 259 (52%) isolated during the study. None of the 193 child household contacts were vaccinated and 47% of them (n=90) isolated during the study period.

**Table 1.**
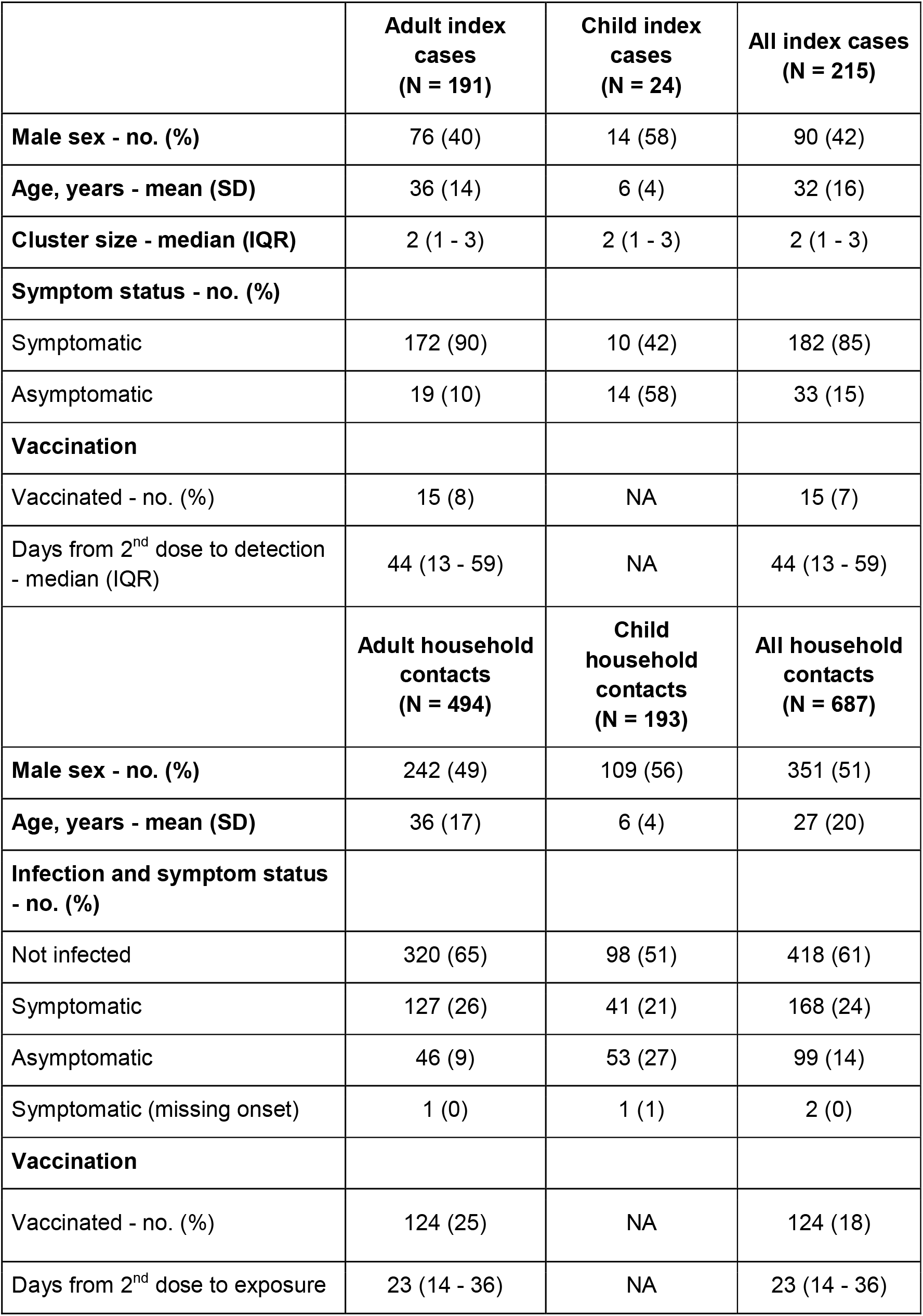
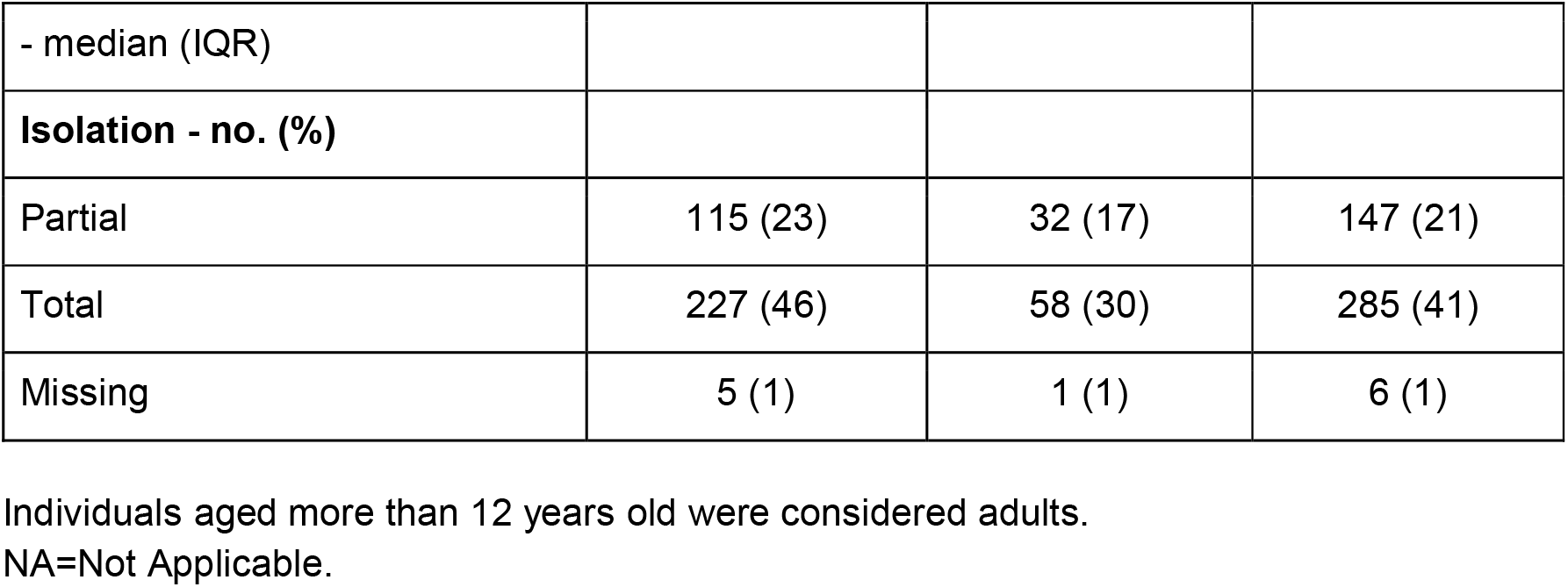
Characteristics of the index cases and household contacts according to their age.

|  | **Adult index cases**<br>**(N = 191)** | **Child index cases**<br>**(N = 24)** | **All index cases**<br>**(N = 215)** |
| --- | --- | --- | --- |
| **Male sex - no. (%)** | 76 (40) | 14 (58) | 90 (42) |
| **Age, years - mean (SD)** | 36 (14) | 6 (4) | 32 (16) |
| **Cluster size - median (IQR)** | 2 (1 - 3) | 2 (1 - 3) | 2 (1 - 3) |
| **Symptom status - no. (%)** |  |  |  |
| Symptomatic | 172 (90) | 10 (42) | 182 (85) |
| Asymptomatic | 19 (10) | 14 (58) | 33 (15) |
| **Vaccination** |  |  |  |
| Vaccinated - no. (%) | 15 (8) | NA | 15 (7) |
| Days from 2<sup>nd</sup> dose to detection - median (IQR) | 44 (13 - 59) | NA | 44 (13 - 59) |
|  | **Adult household contacts**<br>**(N = 494)** | **Child household contacts**<br>**(N = 193)** | **All household contacts**<br>**(N = 687)** |
| **Male sex - no. (%)** | 242 (49) | 109 (56) | 351 (51) |
| **Age, years - mean (SD)** | 36 (17) | 6 (4) | 27 (20) |
| **Infection and symptom status - no. (%)** |  |  |  |
| Not infected | 320 (65) | 98 (51) | 418 (61) |
| Symptomatic | 127 (26) | 41 (21) | 168 (24) |
| Asymptomatic | 46 (9) | 53 (27) | 99 (14) |
| Symptomatic (missing onset) | 1 (0) | 1 (1) | 2 (0) |
| **Vaccination** |  |  |  |
| Vaccinated - no. (%) | 124 (25) | NA | 124 (18) |
| Days from 2<sup>nd</sup> dose to exposure | 23 (14 - 36) | NA | 23 (14 - 36) |
| - median (IQR) |  |  |  |
| **Isolation - no. (%)** |  |  |  |
| Partial | 115 (23) | 32 (17) | 147 (21) |
| Total | 227 (46) | 58 (30) | 285 (41) |
| Missing | 5 (1) | 1 (1) | 6 (1) |

Individuals aged more than 12 years old were considered adults.
NA=Not Applicable.

**Figure 1.**
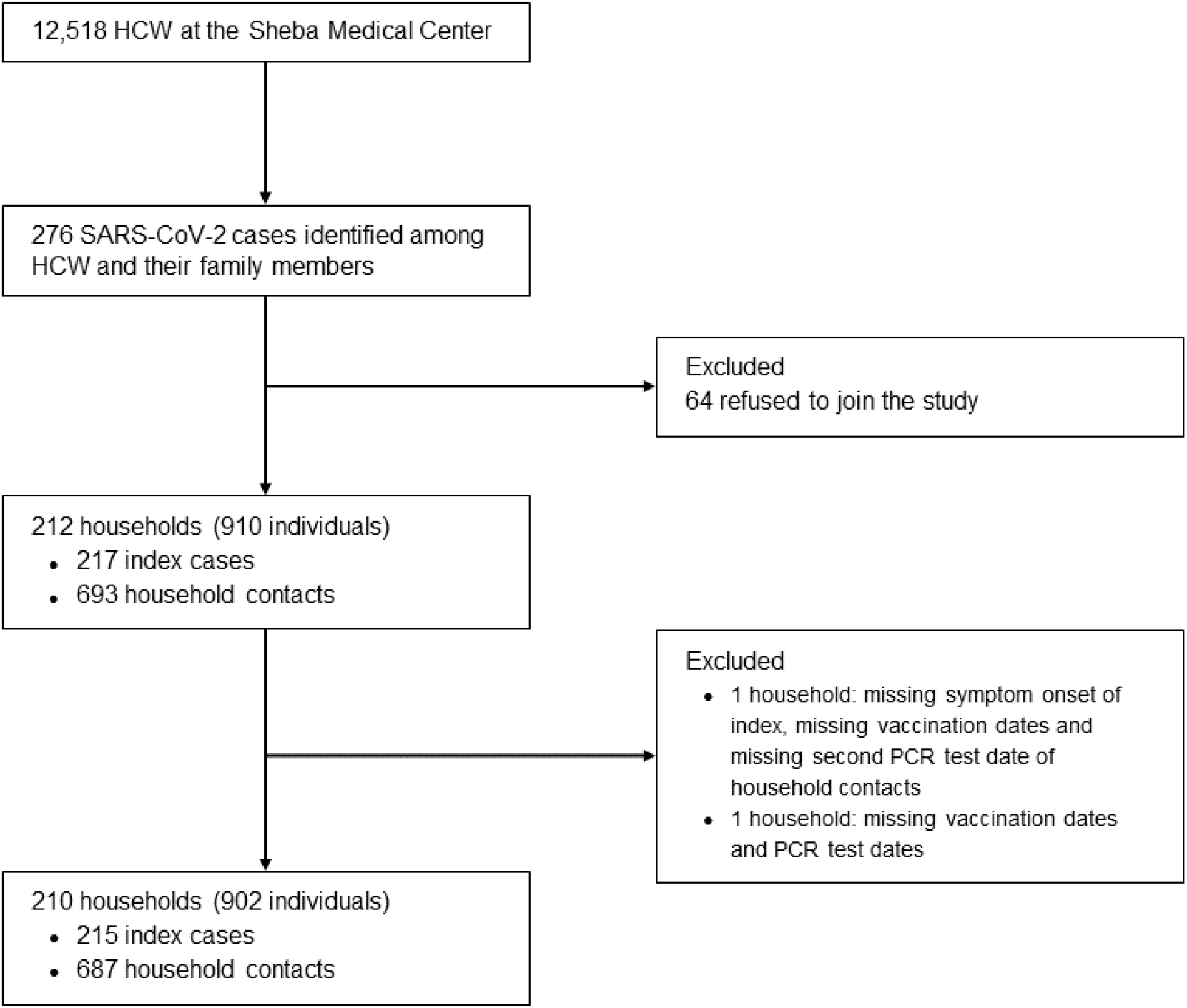
Flow chart of the households included in our analysis.

269 out of 687 (39%) household contacts developed a SARS-CoV-2 infection. Of those, 170 (63%) developed symptoms (Table 1). The secondary attack rate varied with the characteristics of the contacts. Among the 108 adult/teenager contacts who were not vaccinated nor isolated, 81 (75%) were infected by SARS-CoV-2 (Table 2). This proportion dropped to 26% (10 out of 39) among adult/teenager household contacts that were vaccinated but not isolated, 27% (71 out of 259) among adult/teenager household contacts that were isolated but not vaccinated, and 11% (9 out of 83) among adult/teenager household contacts that were vaccinated and isolated. 65% (66 out of 102) of child contacts that were not isolated got infected by SARS-CoV-2. This proportion declined to 32% (29 out of 90) for isolated child contacts. The proportion of asymptomatic cases varied from 26% (46 out of 174) among adult/teenager contact cases to 56% (53 out of 95) among child contact cases.

**Table 2.** Univariate secondary attack rates (SAR) according to the type of contact.

|  | **Infected contacts / Total contacts** | **SAR - %** |
| --- | --- | --- |
| **Contacts** |  |  |
| Adult | 81 / 108 | 75 |
| Vaccinated adult | 71 / 259 | 27 |
| Vaccinated adult | 10 / 39 | 26 |
| Vaccinated + isolated adult | 9 / 83 | 11 |
| Child | 66 / 102 | 65 |
| Isolated child | 29 / 90 | 32 |
| **Index** |  |  |
| Vaccinated | 8 / 43 | 19 |
| Unvaccinated | 261 / 641 | 41 |

The first 6 rows correspond to the SAR of SARS-CoV-2 infection among contacts belonging to the following categories regardless of the vaccination status of the index case: not isolated and not vaccinated adults/teenagers ("Adult"), isolated and not vaccinated adults/teenagers ("Isolated adult"), vaccinated and not isolated adults/teenagers ("Vaccinated adult"), vaccinated and isolated adults/teenagers ("Vaccinated + isolated adult"), not isolated children ("Child"), and isolated children ("Isolated child"). Isolation is missing for one child contact and for 5 adult contacts. The last two rows correspond to the SAR among the household contacts of vaccinated (n=14 households) and unvaccinated index cases (n=195 households). One household was excluded from this analysis because its co-primary cases did not have the same vaccination status.

The secondary attack rate also varied with the vaccination status of the index case regardless of the contacts’ characteristics. Among the 641 household contacts whose index case was unvaccinated, 261 (41%) developed a SARS-CoV-2 infection (Table 2). This proportion dropped to 19% (8 out of 43) among household contacts whose index case was vaccinated. Finally, the secondary attack rate was relatively invariant with household size: 31%, 39%, 32%, and 32% for households of size 2 to 5, respectively (Supplementary Figure 1).

Our statistical model makes it possible to perform a multivariate analysis of the drivers of SARS-CoV-2 transmission in households. We estimate that, relative to adult/teenager contacts who were not vaccinated and did not isolate, the relative risk of being infected was 0·19 (95% CI 0·07-0·40) among adult/teenager household contacts who were vaccinated but did not isolate (Figure 2A). It was 0.11 (95% CI 0·05-0·19) among household contacts who did isolate and were not vaccinated and 0·07 (95% CI 0·03-0·17) among household contacts who were both isolated and vaccinated. Isolation might reduce the risk of infection among vaccinated contacts (93% posterior probability, BF = 14) with a relative risk of 0·39 (95% CI 0·12-1·37). Relative to adult/teenager contacts who were not vaccinated and did not isolate, the relative risk of infection was 0·50 (95% CI 0·32-0·79) for child contacts that did not isolate and 0·16 (95% CI 0·07-0·31) for those that did. We estimate that the risk of transmission from vaccinated cases was 0·22 (95% CI 0·06-0·70) times that of unvaccinated cases (Figure 2B).

**Figure 2.**
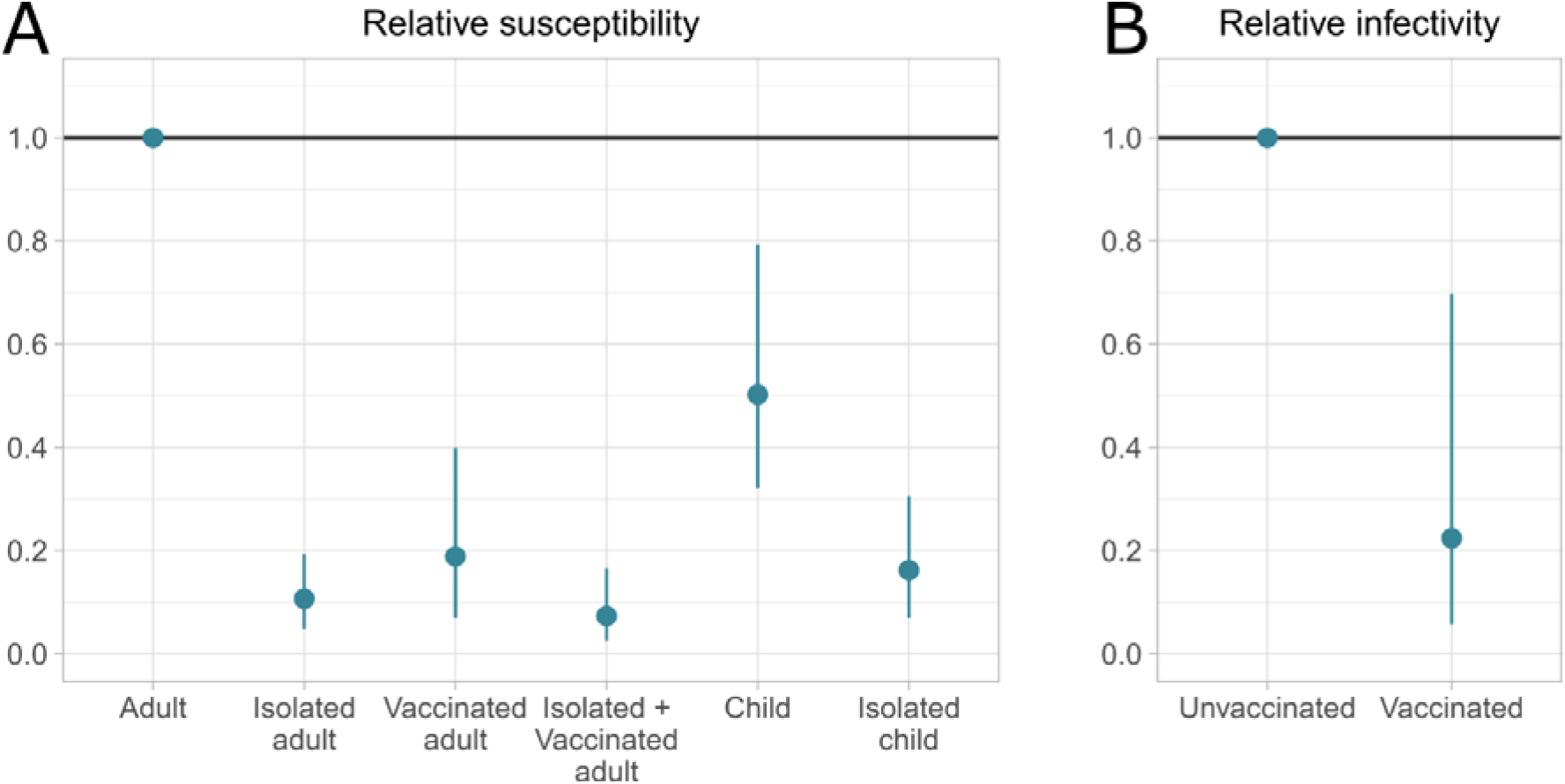
Estimates of SARS-CoV-2 transmission parameters within households. **(A)** Estimated relative susceptibility of isolated and not vaccinated adults/teenagers (“Isolated adult”), vaccinated and not isolated adults/teenagers (“Vaccinated adult”), isolated and vaccinated adults/teenagers (“Isolated + vaccinated adult”), not isolated children (“Child”), and isolated children (“Isolated child”) versus unisolated unvaccinated adults/teenagers (“Adult”). **(B)** Estimated relative infectivity of vaccinated cases compared to unvaccinated cases. The posterior median and its associated 95% Bayesian credible interval are reported.

Overall, we estimate that, in a household of size 4, the probability of SARS-CoV-2 transmission is 59% (95% CI 46-70) between an unvaccinated case and an unvaccinated adult/teenager but 4% (95% CI 1-13) between two vaccinated adults/teenagers (Figure 3). The probability of transmission from an unvaccinated case to a child is 36% (95% CI 26-47). This probability drops to 10% (95% CI 2-27) if the case is vaccinated and to 13% (95% CI 6-23) if the child contact is isolated.

**Figure 3.**
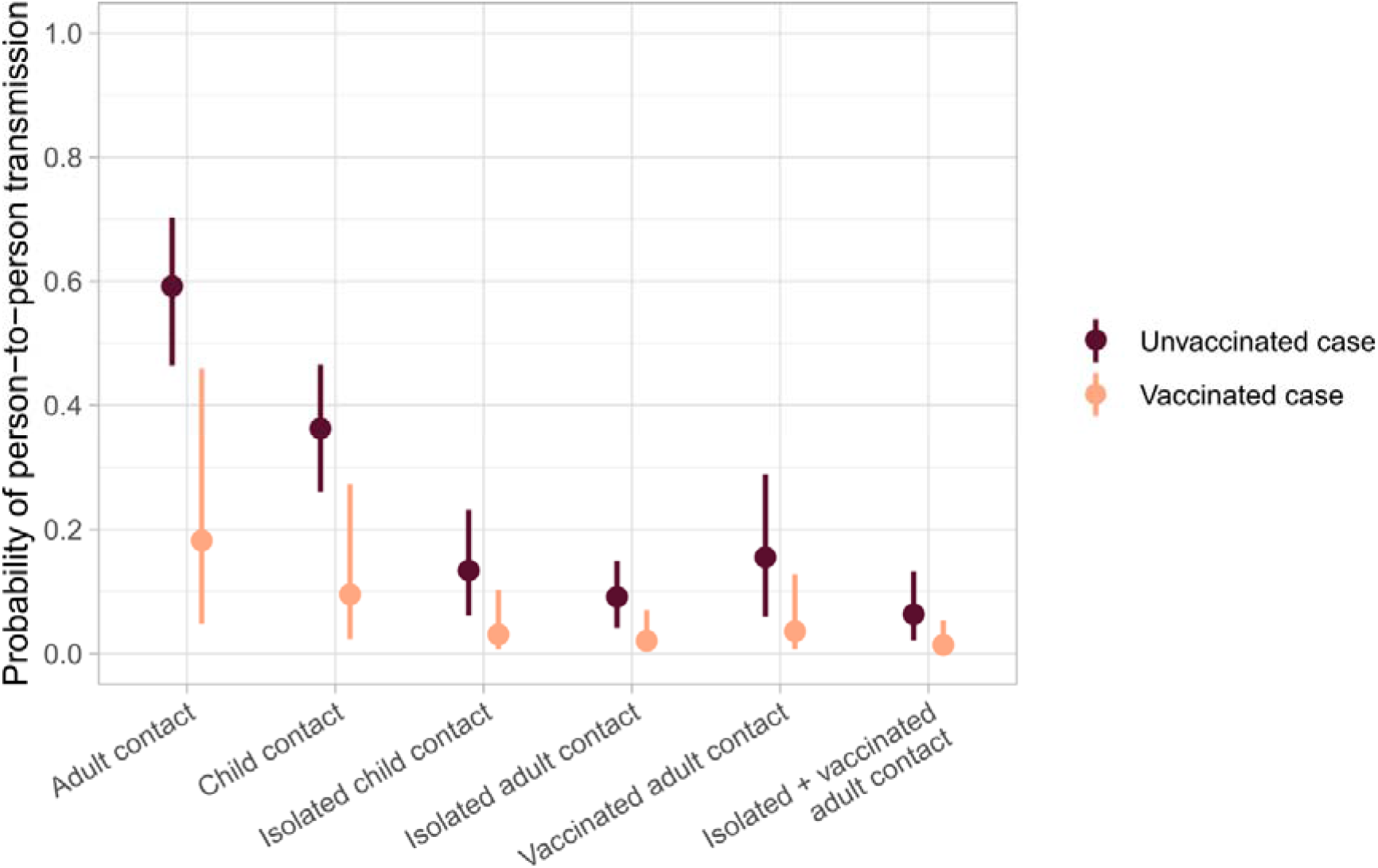
Probability of person-to-person transmission within households according to the characteristics of the case and of the contact. Estimated probability of person-to-person transmission within households of size 4 decomposed by the age, isolation behavior and vaccination status of the contact as well as the vaccination status of the case. “Adult contact” refers to unisolated unvaccinated adult/teenager contacts, “Child contact” refers to not isolated child contacts, “Isolated child contact” refers to isolated child contacts, “Isolated adult contact” refers to isolated and not vaccinated adult/teenager contacts, “Vaccinated adult contact” refers to vaccinated and not isolated adult/teenager contacts, “Isolated + vaccinated adult contacts” refers to isolated vaccinated adult/teenager contacts. The posterior median and its associated 95% Bayesian credible interval are reported.

In general, our estimates of relative susceptibility and relative infectivity were robust to model assumptions (Figure 4). When the analysis was restricted to households in which all contacts performed a PCR test in the ten days following the recruitment of the index case, the relative susceptibility of isolated and vaccinated adult/teenager contacts was slightly higher compared to the baseline scenario. It increased from 0·07 (95% CI 0·03-0·17) to 0·15 (95% CI 0·05-0·35). Consequently, the posterior probability that isolated and vaccinated adult/teenager contacts were less susceptible than vaccinated adult/teenager contacts that did not isolate dropped from 93% to 73% (BF = 2·7), indicating that the estimate of the impact of isolation on vaccinated individuals depends on some of our model assumptions. Relative infectivity and relative susceptibility were slightly sensitive to their prior distribution. When the log-standard deviation increased, estimates were pulled towards lower values.

**Figure 4.**
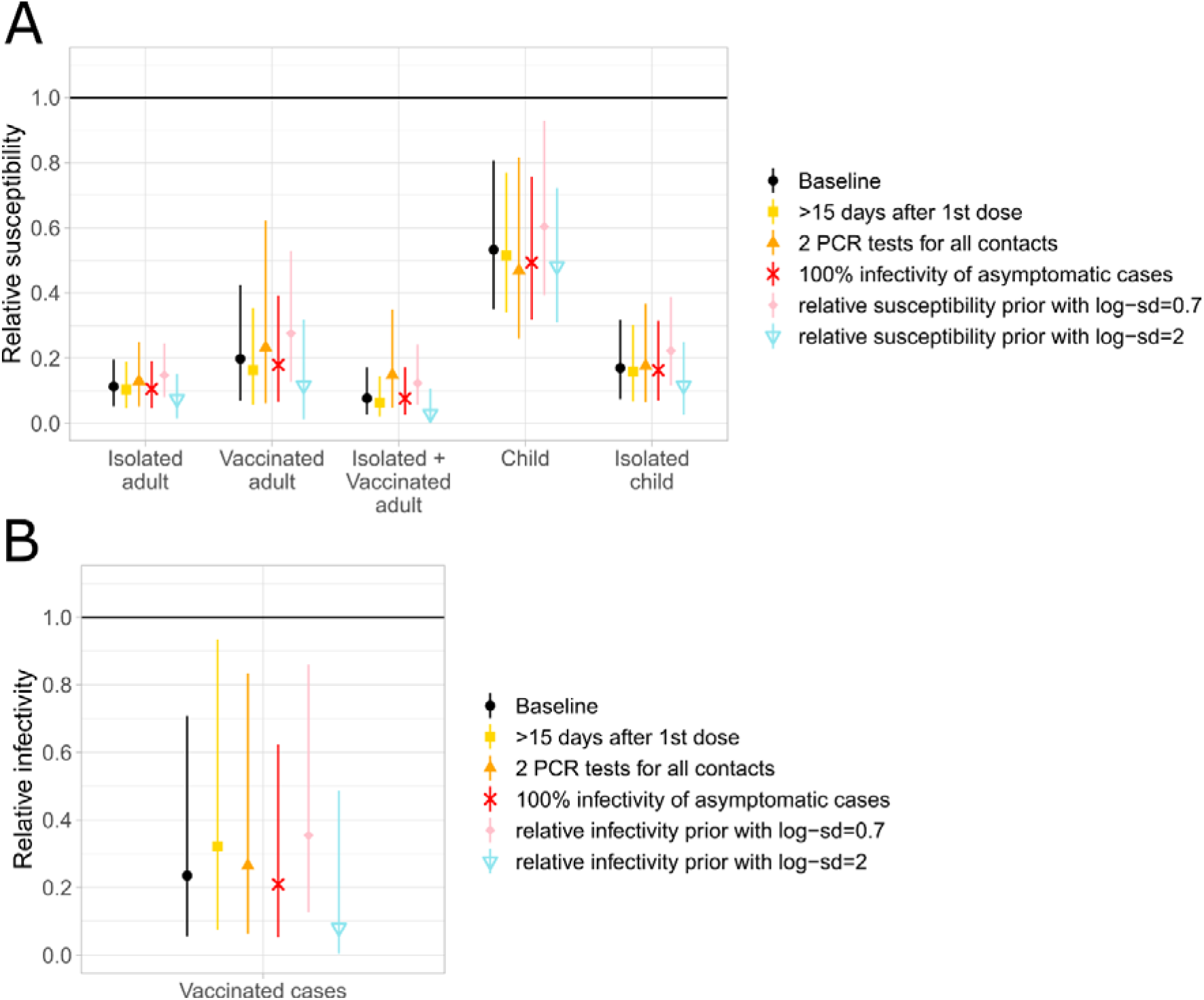
Impact of model assumptions on the estimation of the relative susceptibility and relative infectivity parameters. **(A)** Estimates of the relative susceptibility of household contacts for the baseline and sensitivity analysis scenarios. “Isolated adult” refers to isolated and not vaccinated adults/teenagers, “Vaccinated adult” refers to vaccinated and not isolated adults/teenagers, “Isolated + vaccinated adult” refers to isolated vaccinated adults/teenagers, “Child” refers to not isolated children, and “Isolated child” refers to isolated children. **(B)** Estimates of the relative infectivity of vaccinated cases compared to unvaccinated ones for the baseline and sensitivity analysis scenarios. In the baseline scenario, we assumed that vaccination was effective from 7 days after the 2^nd^ dose, the relative infectivity of asymptomatic cases compared to symptomatic cases was equal to 60% and the log-sd of the relative infectivity and relative susceptibility prior distributions was equal to 1. The posterior median and its associated 95% Bayesian credible interval are reported.

## Discussion

We evaluated the impact of BNT162b2 vaccination on case infectivity and the mitigating effect of age, isolation from the index case and BNT162b2 vaccination on susceptibility to infection in household settings. Our approach accounts for infections in the community, potential tertiary infections within the households, the reduced infectivity of asymptomatic cases, potential misidentification of the index case(s), and varying follow-up periods between households.

In our analysis, the SAR in unvaccinated adult/teenager contacts was estimated at around 75%, which is substantially higher than previous estimates obtained in household settings.^11,14,15,23^ For most of these studies,^14,15,23^ the SAR ranged between 14% and 32% and was estimated when historical lineages were still dominant. In contrast, our study took place when the alpha variant represented up to 90% of transmission events in Israel.^24^ Our higher estimate could be at least partly explained by the fact that the alpha variant is substantially more transmissible than historical lineages.^24–27^ Harris et al.^11^ studied SARS-CoV-2 transmission within households when the alpha variant was dominant in England. Their estimate did not exceed 10% which is likely an under-estimate due to the passive nature of their surveillance.

In agreement with previous reports, we found that children are less susceptible to SARS-CoV-2 infections than adults/teenagers.^14,28,29^ We further estimated that, seven days after their second dose, vaccinated individuals benefit from an 81% reduction in the risk of infection compared to unvaccinated individuals. Consistently with previous studies,^24,30^ we show that BNT162b2 vaccination is highly effective against infection by the alpha variant. In general population studies, vaccine effectiveness for symptomatic infections ranged from 57%^4^ 14 days after the 1^st^ dose to 89%,^4^ and 97%^8^ 7 days after the 2^nd^ dose. For asymptomatic infections, vaccine effectiveness against infection was 79% ten days after the 1^st^ dose,^5^ and 94%^7^ 14 days after the 2^nd^ dose. Our estimate of vaccine effectiveness in household settings is lower than those obtained in the general population. This is consistent with estimates obtained in households by Pritchard et al.^30^ and might in part be explained by the elevated contact rates in households that may favor transmission. Additionally, studies in the general population are less suitable to detect all asymptomatic cases compared to the household setting. This might lead general population studies to over-estimate vaccine efficacy against asymptomatic infections.

To our knowledge, this is the first study estimating the effect of isolation on SARS-CoV-2 transmission in households that are partially vaccinated. We showed that isolation from the index case markedly reduces the overall infection risk in both adult/teenager and child contacts even when considering partial physical distancing measures. We estimated a similar reduction of transmission in adults/teenagers that were vaccinated but not isolated. There was a signal in the data that isolation also benefited vaccinated individuals althought credible intervals were larger and further investigations are required to confirm this finding.

Our study has several limitations. First, household studies such as ours may be affected by multiple sources of bias. On the one hand, we may overestimate the SAR if we are more likely to detect household with multiple cases. On the other hand, we might underestimate it if some asymptomatic, or pauci-symptomatic cases are missed by surveillance. Second, we estimated an important reduction of infectivity in vaccinated cases with 2 doses compared to unvaccinated cases as previously shown.^10,11^ However, this is associated with important uncertainty due to the small number of cases (15 vaccinated index cases, and 19 vaccinated secondary cases). Thus, more data are needed to reduce the size of credible intervals. Third, we assumed an all-or-nothing effect of the vaccine that started seven days after the second dose (or 15 days after the first dose in our sensitivity analysis). In practice, the effect of the vaccine is likely to be progressive, which might push down estimates of efficacy since individuals with early partial protection would be considered as unvaccinated. However, excluding households with the early-vaccinated index cases did not impact our estimates (Supplementary Figure 2). The limited number of households does not make it possible to dissociate early vs full protection conferred by the vaccine nor to investigate the infectivity of children relative to adults/teenagers.

To conclude, vaccination with two doses substantially reduces the risk of transmission and the risk of infection in households. Isolation from the index case while sleeping and eating provides a high level of protection to unvaccinated household members, whether they are adults/teenagers or children. Household contacts of COVID-19 patients should ideally isolate, or at least refrain from significant contacts with household cases. This may also be the case for vaccinated household members although larger studies are required to confirm this finding.

## Data Availability

Data and code will be published online upon publication.

## Acknowledgments

We would like to acknowledge the Infection Prevention & Control Unit team for their devoted work, the extensive epidemiologic investigations and contact tracing from which the data was derived. We thank the Sheba management for their support of this study.

## Author contributions

ML, M Gilboa, SC, and GRY conceived the study. M Gilboa, TG, M Goldenfeld, and LM collected the data. ML, and M Gilboa performed the analyses. ML, M Gilboa, and SC wrote the first draft. All authors contributed to revisions of the manuscript.

## Data and materials availability

Data and code will be published online upon publication.

