## Supplementary Materials for "Impact of BNT162b2 vaccination and isolation on SARS-CoV-2 transmission in Israeli households: an observational study"

**Table of Contents**


**Model of SARS-CoV-2 transmission dynamics in households** 2
- Overview 2
- Transmission within households 2
- Instantaneous risk of infection of a household member 3
- Likelihood function 3

**Inference framework** 3
- Priors 4
- Algorithm 4
- Implementation 4

**Model adequacy** 4

**Parameter estimates** 6

**Sensitivity analysis** 7
- Secondary attack rates 7
- Prior distributions of the relative infectivity and relative susceptibility parameters 8
- Vaccination definition 8
  - Early vaccination 8
  - Effective vaccination >15 days after the 1st dose 9
- Households where all contacts performed a PCR test in the ten days following the detection of the index case 10

**References** 14

### 1. Model of SARS-CoV-2 transmission dynamics in households

#### a. Overview

We developed a statistical model describing SARS-CoV-2 transmission within households that accounts for tertiary infections (household members infected by a household case who is not the index case), infection events in the community, household size, and varying follow up periods between households.

For an individual $i$ in household $k$, data consist in a vector $(a_i, s_i, d_i, v_i, m_i, t_{end})$ where $a_i$ indicates whether $i$ is an adult/teenager above 12 years old or a child, $s_i$ is the infection status of individual $i$ (symptomatic infection, asymptomatic infection or not infected), $d_i$ is the symptom onset date for symptomatic cases or the date of the first positive RT-qPCR test for asymptomatic cases, $v_i$ is the vaccination status of $i$, $m_i$ indicates whether $i$ isolated from the index case when applicable, and $t_{end}$ is the end of the follow up period of household $k$. For each confirmed case, we augmented their observed data with their unobserved date of infection. Infection dates were defined as continuous time to ensure the ordering of infection events within households. Time 0 corresponds to the first infection time in each household. Within household $k$, $I_k$ denotes the list of SARS-CoV-2 cases and $S_k$ denotes the list of susceptibles.

Model parameters were estimated in a Bayesian Markov Chain Monte Carlo (MCMC) framework [1].

#### b. Transmission within households

If we consider infector $i$ and infectee $j$ in household $k$ of size $n$, the instantaneous hazard that $i$ infects $j$ at time $t$ is:

$$h_{i \to j}(t) = \frac{\beta}{n^{\delta}} \mu_{sus}(a_j, m_j, v_j) \, \mu_{inf}(v_i) \, \mu_{asymp}(s_i) \, f\left(t - \xi_i \,\middle|\, d_i, s_i\right)$$

where:

- $\beta / n^{\delta}$ models the dependency between the transmission rate and the household size $n$.

- $\mu_{sus}(a_j, m_j, v_j)$ is the relative susceptibility of recipient $j$ according to their age, vaccination status and isolation status. We define 5 categories of contacts (isolated and not vaccinated adults/teenagers, vaccinated and not isolated adults/teenagers, isolated and vaccinated adults/teenagers, isolated children, not isolated children) that are compared to adults/teenagers who did not isolate and were not vaccinated. For the reference group, $\mu_{sus}(a_j, m_j, v_j) = 1$.

- $\mu_{inf}(v_i)$ is the relative infectivity of infector $i$ according to its vaccination status.

$$\mu_{inf}(v_i) = \begin{cases} 1 & \text{if } v_i = 0 \\ \pi_{inf} & \text{if } v_i = 1 \end{cases}$$

- $\mu_{asymp}(s_i)$ is the relative infectivity of infector $i$ whether $i$ is symptomatic or asymptomatic.

$$\mu_{asymp}(s_i) = \begin{cases} 1 & \text{if } s_i = 1 \\ \pi_{asymp} & \text{if } s_i = 0 \end{cases}$$

  In the baseline scenario, we assumed a 40% reduction of the infectivity of asymptomatic cases compared to symptomatic cases ($\pi_{asymp} = 0.6$) as estimated by Byambasuren et al., 2020 [2]. In the sensitivity analysis, we explored the impact of similar infectivity levels between symptomatic and asymptomatic cases ($\pi_{asymp} = 1.0$) on the estimation of the relative susceptibility and relative infectivity parameters.

- $f(\Delta t \mid d, s)$ is the density of the generation time defined as the distribution of the interval between the infection time $\xi_i$ of the infector $i$ and the infection time $\xi_j$ of the recipient $j$ conditioned on the symptom onset or date of detection, and the presence of symptoms of $i$.
  For symptomatic cases, $f$ is derived from the corrected infectivity profile estimated by Ashcroft et al., 2020 [3], a shifted $\Gamma$ distribution with shape=97.2, rate=3.7 and shift=25.6. The shift of the distribution corresponds to the symptom onset of $i$. According to McAloon et al., 2020, less than 2% of the symptomatic cases develop symptoms over the three days following their infection [4]. We assumed that incubation periods are of 3 days minimum and that the infectious period starts 3 days before the symptom onset, independent of the duration of the incubation period.
  For asymptomatic infectors, $f$ is derived from the same estimate of the infectivity profile so that infectors are infectious starting from 2 days after their infection time and their infectivity peaks approximately 5 days after their infection.

#### c. Instantaneous risk of infection of a household member

The risk of infection of individual $j$ in household $k$ at time $\xi$ is the sum of the hazard of infection within the community and the hazards of infection by infected household members:

$$\lambda_{j,k}(\xi) = \alpha + \sum_{i \in I_k \{\xi_i < \xi\}} h_{i \to j,k}(\xi)$$

where $\alpha$ is the instantaneous risk of infection in the community. It is assumed constant over the follow-up of households and the entire period of the study.

#### d. Likelihood function

Denote $\theta$ the vector of the transmission model parameters. The likelihood of the transmission process within the household conditional on the first date of infection $\xi_1$ in the household is:

$$P(\xi \mid \theta) = \prod_{i \in I} f\left(d_i - \xi_i\right) \prod_{i \in I - \{1\}} \lambda_i(\xi_i)\, e^{-\int_{\xi_1}^{\xi_i} \lambda_i(u)\, \mathrm{d}u} \prod_{j \in S} e^{-\int_{\xi_1}^{t_{end}} \lambda_j(u)\, \mathrm{d}u}$$

where $f(d_i - \xi_i)$ is the density of the incubation period for symptomatic cases or the density of the RT-qPCR detection period after infection for asymptomatic cases. The distribution of the incubation period was defined as a truncated log-normal distribution with log-mean=1.63 and log-sd=0.25 as estimated by McAloon et al., 2020 [2,4]. As previously mentioned, less than 2% of the symptomatic cases develop symptoms over the three days following their infection according to this distribution. We assumed that the incubation period lasts at least 3 days and does not exceed 30 days. For the RT-qPCR detection period, we assumed a Uniform(0,10) distribution.

For the incubation period and the infectivity profile, we used distributions that were estimated on data from the historical lineages. Over the study period, up to 80% of the infections were caused by the alpha variant in Israel [5]. Yet, there is little knowledge on the mechanisms underlying the rapid spread of the alpha variant at the individual level. Modelling and phylodynamic studies support the hypothesis of its higher transmissibility compared to the historical lineages. The results concerning potentially shorter generation time and longer infectious period are less clear [6–8]. Since participants were not screened for the variants, we assumed that the infectivity profile and the incubation period remained unchanged between the alpha variant and the historical lineages.

### 2. Inference framework

We used a data augmentation MCMC approach to explore the joint posterior distribution of model parameters and the augmented dates of infection.

#### a. Priors

We choose a Uniform(0,1) prior distribution for the hazard of infection within the community $\alpha$ and a Uniform(0,5) prior distribution for the per capita transmission rate within households $\beta$. For the dependency between the transmission rate and the household size, the prior distribution for $\delta$ was a Uniform(-3,3) distribution.

For the relative susceptibility of the different categories of contacts $\mu_{sus}$ and the relative infectivity of vaccinated cases $\mu_{inf}$, a log-normal prior with log-mean=0 and log-sd=1 was used. We investigated the impact of the log-sd on parameter estimation in a sensitivity analysis (Supplementary Materials 5.b.).

#### b. Algorithm

Parameters were updated using a Metropolis-Hastings algorithm.

Data augmentation was performed at each iteration of the MCMC chain. After the update of the parameters, the infection times of all COVID-19 cases were updated. For symptomatic cases, the incubation period was sampled from the distribution mentioned above and the infection date was obtained as the difference of the symptom onset and the incubation period. For asymptomatic cases, we chose a conservative scenario according to which asymptomatic cases could have been infected up to ten days prior to their first positive RT-qPCR test independent of the Ct value of the test.

#### c. Implementation

The data augmentation Markov Chain Monte Carlo sampling algorithm was implemented in C++. Chains were run for 100,000 iterations and one out of 10 iterations were recorded. Marginal posteriors were sampled from MCMC chains after discarding a burn-in of 10,000 steps. Convergence was inspected visually.

### 3. Model adequacy

To assess the adequation of the model to the data, we performed a simulation study. Using an agent-based model, we simulated 2,000 data sets with a structure identical to that of the observed data (household size, age, vaccination and isolation status of the household members, symptom status of the index case, proportion of secondary asymptomatic cases, and follow-up period) with parameters equal to samples from their joint posterior distribution. We compared the observed secondary attack rate (SAR) to the one expected under the model estimates. There was a good agreement between the observed and expected SAR for households of size 2 to 5.

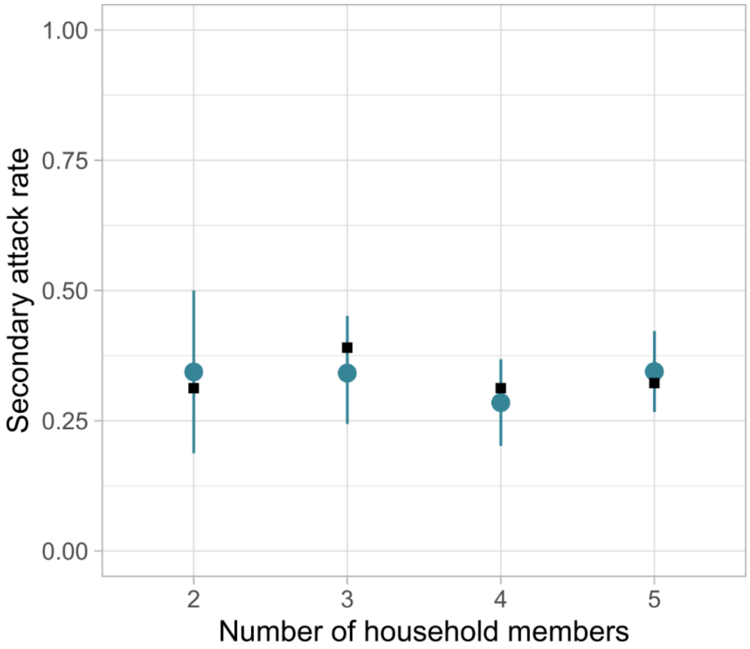


**Supplementary Figure 1. Observed and expected secondary attack rates under the model estimates for households of size 2 to 5.**
Black squares correspond to the observed SAR per household size. The mean and 95% CI are reported.

We also compared the observed and expected distributions of the number of infected individuals per household size. The observed values fall within the 95% CI of the expected values except for the households of size 4 with 2 infected individuals. There is a good agreement between the observed and expected distributions.

**Supplementary Table 1. Comparison of the observed and expected distributions of the number of infected individuals for households of size 2 to 5.**
The mean and the 95% CI are reported.

| Household size | Distribution | Number of infected individuals per household | | | | |
|---|---|---|---|---|---|---|
| | | 1 | 2 | 3 | 4 | 5 |
| 2 | Observed | 22 | 10 | | | |
| 2 | Expected | 21 (16-26) | 11 (6-16) | | | |
| 3 | Observed | 21 | 9 | 12 | | |
| 3 | Expected | 20.5 (14-27) | 13.5 (8-20) | 8 (4-13) | | |
| 4 | Observed | 25 | 9 | 8 | 7 | |
| 4 | Expected | 22.2 (16-29) | 15.6 (9-22) | 7.8 (3-13) | 3.4 (1-7) | |
| 5 | Observed | 19 | 9 | 10 | 2 | 6 |
| 5 | Expected | 13.4 (8-19) | 13.8 (8-20) | 9.7 (5-15) | 6.1 (2-11) | 3 .1 (1-6) |

### 4. Parameter estimates

Supplementary Table 2 compiles the median and 95% CI of the relative susceptibility and relative infectivity estimates that are presented in Figures 2 and 4. Supplementary Table 3 compiles the median and 95% CI of the person-to-person risk of infection represented in Figure 3.

**Supplementary Table 2. Estimates of the relative susceptibility of household contacts and relative infectivity of cases in the baseline scenario and in the sensitivity analysis.**
The median and the 95% CI are reported. In the >15 days after 1st dose scenario, we assumed that vaccination is effective from 15 days after the 1st injection. In the 2 PCR tests for all contacts, we restricted the analysis to the households where all household members performed a PCR test in the 10 days following the detection of the index case. In the 100% infectivity of asymptomatic cases, we assumed that symptomatic and asymptomatic cases have the same level of infectivity. In the relative susceptibility prior with log-sd=0.7 scenario, we used a log-sd=0.7 for the prior of the relative susceptibility parameters. In the relative infectivity prior with log-sd=0.7 scenario, we used a log-sd=0.7 for the prior of the relative infectivity of vaccinated cases compared to unvaccinated cases parameter. In the baseline scenario, we assumed that vaccination was effective from 7 days after the 2nd dose, the relative infectivity of asymptomatic cases compared to symptomatic cases was equal to 60% and the log-sd of the relative infectivity and relative susceptibility prior distributions was equal to 1. The posterior median and its associated 95% Bayesian credible interval are reported.

| | **Baseline scenario** | **>15 days after 1st dose** | **2 PCR tests for all contacts** | **100% infectivity of asymptomatic cases** | **Relative susceptibility prior with log-sd=0.7** | **Relative susceptibility prior with log-sd=2** |
|---|---|---|---|---|---|---|
| **Relative susceptibility** | | | | | | |
| Isolated and not vaccinated adult/teenager | 0.11 (0.05-0.19) | 0.09 (0.04-0.17) | 0.13 (0.05-0.25) | 0.1 (0.05-0.19) | 0.15 (0.08-0.24) | 0.07 (0.01-0.14) |
| Vaccinated and not isolated adult/teenager | 0.19 (0.07-0.4) | 0.15 (0.05-0.32) | 0.23 (0.06-0.62) | 0.18 (0.06-0.38) | 0.27 (0.12-0.53) | 0.11 (0.01-0.3) |
| Isolated + vaccinated adult/teenager | 0.07 (0.03-0.17) | 0.06 (0.02-0.13) | 0.15 (0.05-0.35) | 0.08 (0.03-0.17) | 0.12 (0.06-0.24) | 0.03 (0-0.1) |
| Not isolated child | 0.5 (0.32-0.79) | 0.46 (0.29-0.71) | 0.46 (0.26-0.8) | 0.49 (0.31-0.77) | 0.6 (0.38-0.93) | 0.43 (0.27-0.68) |
| Isolated child | 0.16 (0.07-0.31) | 0.14 (0.06-0.27) | 0.17 (0.06-0.37) | 0.16 (0.07-0.31) | 0.22 (0.11-0.39) | 0.11 (0.03-0.23) |
| **Relative infectivity** | | | | | | |
| Vaccinated case | 0.22 (0.06-0.7) | 0.3 (0.07-0.87) | 0.26 (0.06-0.84) | 0.21 (0.05-0.64) | 0.35 (0.12-0.85) | 0.08 (0-0.47) |

**Supplementary Table 3. Estimates of the transmission probability from vaccinated adult/teenager cases and unvaccinated cases to the different categories of contacts within households.**
Probabilities are reported in percentage with their 95% CI.

| Contact | Vaccinated adult/teenager case | Unvaccinated case |
|---|---|---|
| **Not isolated and not vaccinated adult/teenager** | 18.2 (4.8-45.9) | 59.2 (46.4-70.2) |
| **Isolated and not vaccinated adult/teenager** | 2.1 (0.4-7) | 9.2 (4.2-15) |
| **Isolated + vaccinated adult/teenager** | 1.4 (0.3-5.3) | 6.4 (2.2-13.3) |
| **Vaccinated and not isolated adult/teenager** | 3.6 (0.7-12.8) | 15.5 (5.9-28.9) |
| **Not isolated child** | 9.6 (2.4-27.3) | 36.2 (26.1-46.5) |
| **Isolated child** | 3.1 (0.7-10.3) | 13.4 (6.2-23.1) |

### 5. Sensitivity analysis

#### a. Secondary attack rates

In the baseline scenario, we calculated the SAR according to the type of contact for all households regardless of the number of identified index cases. In Supplementary Table 4, we restricted the calculation of the SAR to households where a single index case was identified (n=206). There was barely an impact on the SAR.

**Supplementary Table 4. Univariate secondary attack rates according to the type of contact restricted to households where a single index case was identified.**

| | Infected contacts / Total contacts | SAR - % |
|---|---|---|
| **Contacts** | | |
| Not isolated and not vaccinated adult/teenager | 78 / 103 | 76 |
| Isolated and not vaccinated adult/teenager | 71 / 257 | 28 |
| Vaccinated and not isolated adult/teenager | 10 / 39 | 26 |
| Vaccinated and isolated adult/teenager | 9 / 81 | 11 |
| Not isolated child | 64 / 97 | 66 |
| Isolated child | 29 / 89 | 33 |
| **Index** | | |
| Vaccinated | 8 / 43 | 19 |
| Unvaccinated | 256 / 629 | 41 |

#### b. Prior distributions of the relative infectivity and relative susceptibility parameters

The log-sd of the relative susceptibility and relative infectivity parameters modifies the value range that is likely to be explored. In the baseline scenario, we used a log-sd=1.0 for both the relative infectivity and relative susceptibility parameters which corresponds to a relatively large 95% interval spanning from 0.14 to 7.1. With log-sd=0.7, the interval considerably shrinks around 1 and with log-sd=2 it spans from 0.02 to 50.4.

| **Log-sd of the prior** | **2.5% percentile** | **97.5% percentile** |
|---|---|---|
| **0.7** | 0.25 | 3.9 |
| **1** | 0.14 | 7.1 |
| **2** | 0.02 | 50.4 |

#### c. Vaccination definition

##### i. Early vaccination

To determine the impact of early vaccinated cases on our estimates, we restricted the analysis to the households where the index case was either not vaccinated or infected >7days after the 2nd dose, i.e., this analysis did not contain households where the index was vaccinated but infected before the vaccine was considered effective. The estimations are represented in Supplementary Figure 2 and the values are gathered in Supplementary Table 2. Compared to the baseline scenario, the relative susceptibility parameters were not impacted. However, the estimation of the relative infectivity parameter is less precise with an upper bound of the 95% CI passing from 0.71 to 0.92.

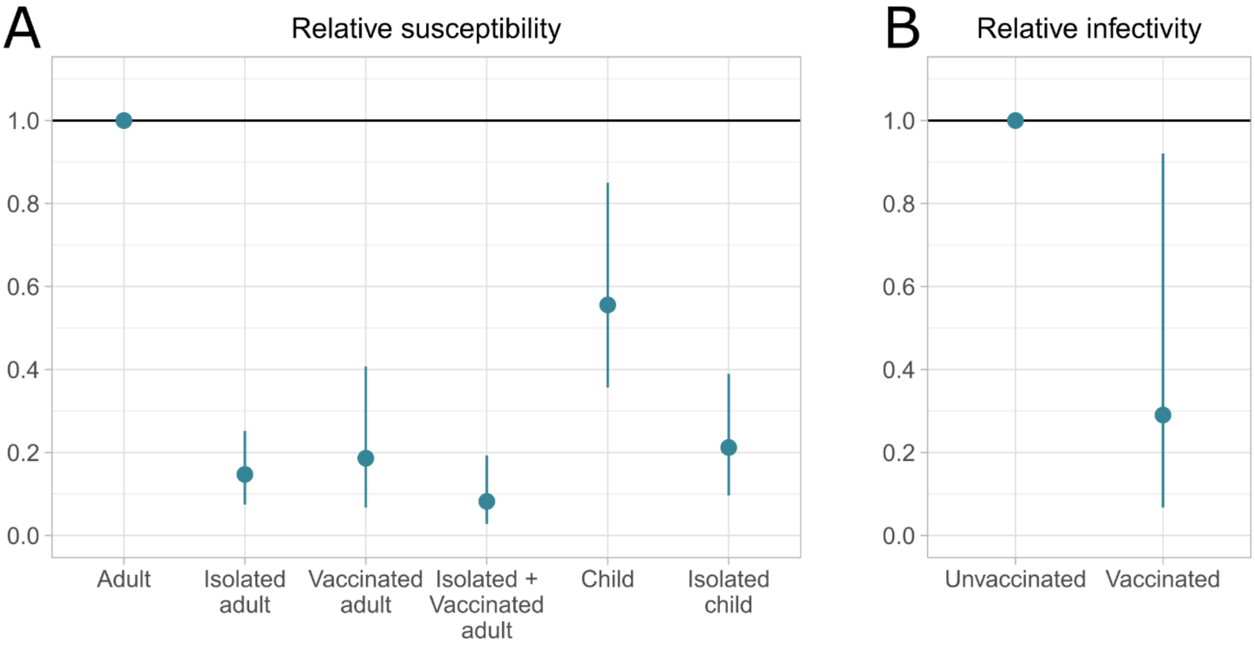


**Supplementary Figure 2. Estimates of the SARS-CoV-2 transmission parameters within households where the index case was not early vaccinated.**
**(A)** Estimated relative susceptibility of isolated and not vaccinated adults/teenagers, vaccinated and not isolated adults/teenagers, isolated and vaccinated adults/teenagers, not isolated children, and isolated children versus not isolated nor vaccinated adults/teenagers. **(B)** Estimated relative infectivity of vaccinated cases compared to unvaccinated cases. The median estimate and its associated 95% Bayesian credibility interval are reported.

**Supplementary Table 5. Estimates of the SARS-CoV-2 transmission parameters within households where the index case was not early vaccinated.**
The median and the 95% CI are reported.

| **Relative susceptibility** | **Baseline scenario** | **Household with no early vaccinated index case** |
|---|---|---|
| Isolated and not vaccinated adult/teenager | 0.11 (0.05-0.2) | 0.14 (0.06-0.25) |
| Vaccinated and not isolated adult/teenager | 0.2 (0.07-0.42) | 0.18 (0.06-0.4) |
| Isolated and vaccinated adult/teenager | 0.08 (0.03-0.17) | 0.08 (0.03-0.19) |
| Not isolated child | 0.53 (0.35-0.81) | 0.56 (0.34-0.94) |
| Isolated child | 0.17 (0.07-0.32) | 0.21 (0.09-0.4) |
| **Relative infectivity - median (95% CI)** | | |
| Vaccinated | 0.23 (0.05-0.71) | 0.24 (0.06-0.75) |

#### ii. Effective vaccination >15 days after the 1st dose

In a sensitivity analysis, we tested how a different definition of effective vaccination (>15 days after the 1st dose or >7 days after the 2nd dose) affects parameter estimates. There are 3 additional vaccinated index cases and 16 additional vaccinated adult/teenager contacts in the 1 dose scenario (sensitivity analysis) compared to the 2 doses scenario (baseline). The 1 dose scenario slightly increases the share of vaccinated adult/teenager index cases and vaccinated adult/teenager contacts, but it did not impact parameter estimates (Figure 4).

**Supplementary Table 5. Vaccination status of the adult/teenager index cases and adult/teenager household contacts according to the definition of effective vaccination.**
3 individuals were exposed to the index case more than 15 days after they received the 1st vaccine dose but did not remember the exact date. 1 index case and 4 household contacts were detected or exposed about 15 days after they received the 1st vaccine dose. These individuals were considered as not vaccinated.

| | **>15 days after the 1st dose (sensitivity analysis)** | **>7 days after the 2nd dose (baseline)** |
|---|---|---|
| **Adult/teenager index cases (N = 191)** | | |
| Vaccinated - no. (%) | 18 (9) | 15 (8) |
| Days from 1st dose to detection - median (IQR) | 42 (29.25 - 78.75) | 44 (13 - 59) |
| Missing vaccination date - no. (%) | 1 (1) | 0 (0) |
| **Adult/teenager contacts (N = 494)** | | |
| Vaccinated - no. (%) | 140 (28) | 124 (25) |

| | | |
|---|---|---|
| Days from 1st dose to exposure - median (IQR) | 42 (33 - 56) | 23 (14 - 36) |
| Missing vaccination date - no. (%) | 4 (1) | 0 (0) |

#### d. Households where all contacts performed a PCR test in the ten days following the detection of the index case

In the baseline scenario, household contacts who did not report symptoms and did not perform a PCR test in the ten days following the detection of the index case were considered not infected over their follow-up (n=126). In a sensitivity analysis, we verified the robustness of our estimates to this hypothesis by removing all the households with at least one contact whose outcome was not confirmed which restricted the analysis to 141 households (Supplementary Figure 3). There were 145 index cases, including 4 co-primary cases, and 429 household contacts, among whom 212 (49%) developed a SARS-CoV-2 infection (Supplementary Table 6). This is slightly higher than in the 39% in the baseline scenario. The characteristics of the index cases are remarkably similar to the baseline scenario except for the median time from the 2$^{nd}$ dose to detection. It is equal to 44 (IQR 13-59) in the baseline scenario and to 56 (IQR 33-62) in the sensitivity analysis. The characteristics of the household contacts are also remarkably similar, notably the proportion of asymptomatic cases that is higher among children (28%) compared to adults/teenagers (11%). However, the median time from the 2nd dose to exposure is slightly higher in the sensitivity (29, IQR 14-46) compared to the baseline scenario (23, IQR 14-36).

Compared to the baseline scenario, the univariate SAR are higher in all contact categories except for the vaccinated adults/teenagers (Supplementary Table 7). The SAR in households with vaccinated index case(s) is equal to 3% in the sensitivity analysis but there are only 29 household contacts. There is low statistical power to precisely estimate the reduction of infectivity in vaccinated index cases compared to unvaccinated index cases which explains why the 95% CI is larger in the sensitivity analysis.

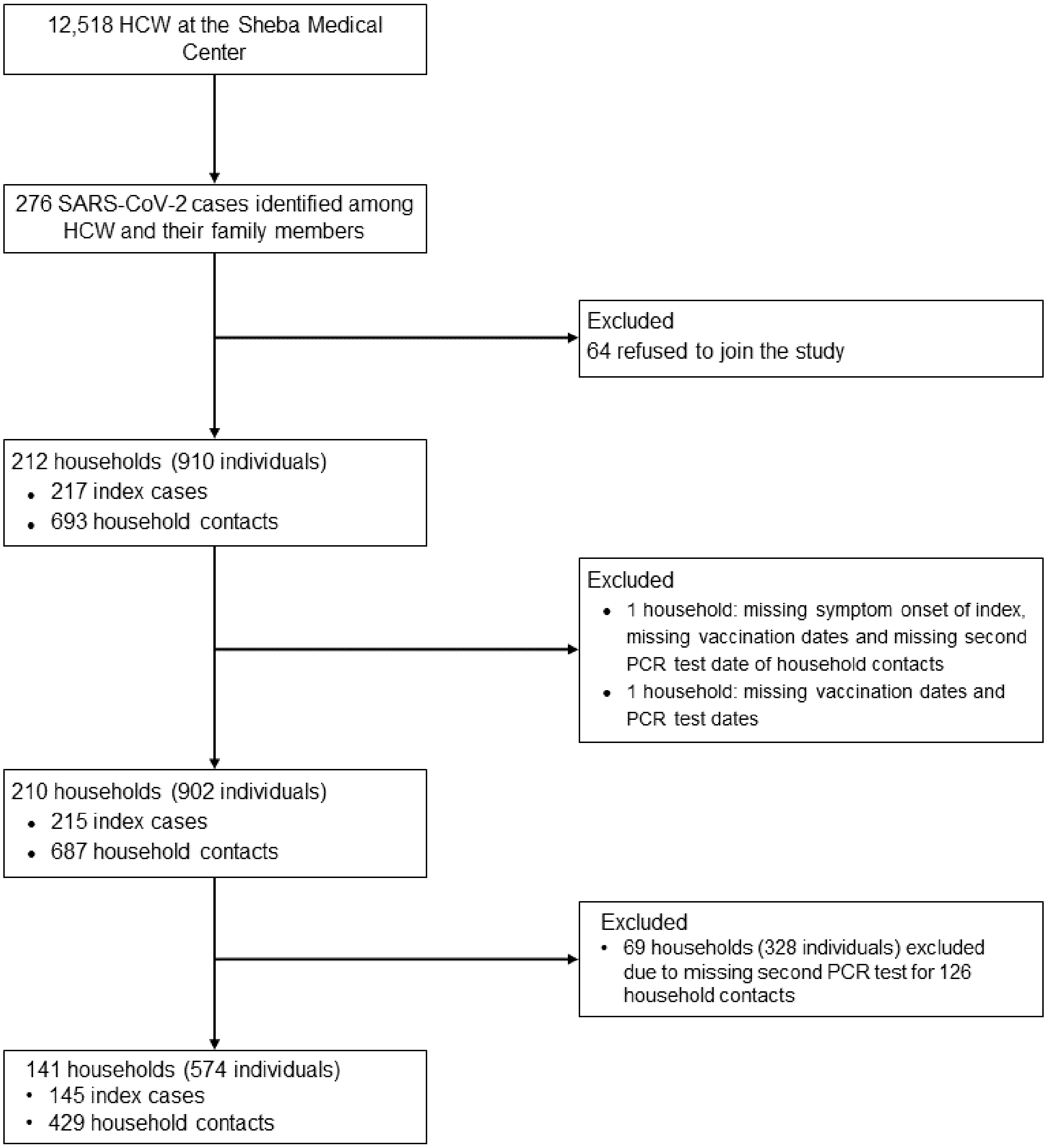


**Supplementary Figure 3. Flow chart of the households included in the sensitivity analysis.**

**Supplementary Table 6. Characteristics of the index cases and household contacts according to their age for households where all contacts had a PCR test in the ten days following the detection of the index case.**

| | **Adult/teenager index cases (N = 139)** | **Child index cases (N = 14)** | **All index cases (N = 153)** |
|---|---|---|---|
| **Male sex - no. (%)** | 60 (43) | 8 (57) | 68 (44) |
| **Age, years - mean (SD)** | 38 (15) | 7 (3) | 35 (16) |
| **Cluster size - median (IQR)** | 2 (1 - 3) | 3 (1.25 - 4) | 2 (1 - 3) |
| **Symptom status - no. (%)** | | | |
| Symptomatic | 123 (88) | 5 (36) | 128 (84) |
| Asymptomatic | 16 (12) | 9 (64) | 25 (16) |
| **Vaccination** | | | |
| Vaccinated - no. (%) | 11 (8) | - | 11 (7) |
| Days from 2nd dose to detection - median (IQR) | 56 (33 - 62) | - | 56 (33 - 62) |
| | **Adult/teenager household contacts (N = 344)** | **Child household contacts (N = 134)** | **All household contacts (N = 478)** |
| **Male sex - no. (%)** | 163 (47) | 73 (54) | 236 (49) |
| **Age, years - mean (SD)** | 35 (17) | 6 (3) | 27 (20) |
| **Infection and symptom status - no. (%)** | | | |
| Not infected | 200 (58) | 66 (49) | 266 (56) |
| Symptomatic | 105 (31) | 30 (22) | 135 (28) |
| Asymptomatic | 38 (11) | 38 (28) | 76 (16) |
| Symptomatic (missing onset) | 1 (0) | 0 (0) | 1 (0) |
| **Vaccination** | | | |
| Vaccinated - no. (%) | 69 (20) | - | 69 (14) |
| Days from 2nd dose to exposure - median (IQR) | 29 (14 - 46) | - | 29 (14 - 46) |
| Missing vaccination date - no. (%) | 0 (0) | - | 0 (0) |
| **Isolation - no. (%)** | | | |
| Partial | 94 (27) | 22 (16) | 116 (24) |
| Total | 143 (42) | 41 (31) | 184 (38) |
| Missing | 4 (1) | 1 (1) | 5 (1) |

**Supplementary Table 7. Univariate secondary attack rates according to the type of contact restricted to households where all contacts had a PCR test in the ten days following the detection of the index case.**

| | **Infected contacts / Total contacts** | **SAR - %** |
|---|---|---|
| **Contacts** | | |
| Not isolated and not vaccinated adult/teenager | 67 / 84 | 80 |
| Isolated and not vaccinated adult/teenager | 62 / 189 | 33 |
| Vaccinated and not isolated adult/teenager | 4 / 19 | 21 |
| Vaccinated and isolated adult/teenager | 9 /48 | 19 |
| Not isolated child | 46 / 70 | 66 |
| Isolated child | 22 / 63 | 35 |
| **Index** | | |
| Vaccinated | 1 / 29 | 3 |
| Unvaccinated | 211 / 449 | 47 |
